# The Sepsis ImmunoScore Predicts Sepsis, Mortality, and Deterioration Better than Clinical Scores and Widely Available Biomarkers

**DOI:** 10.1101/2025.10.01.25337117

**Authors:** Gregory L. Watson, Lincoln C. Updike, Carlos G. López-Espina, Akhil Bhargava, Lee A. Schmalz, Shah Khan, Dennys S. Urdiales, Matthew D. Sims, Ashok V. Palagiri, Adrian D. Haimovich, Alon Dagan, Benjamin P. Davis, Karen C. White, Paul A. Gurbel, Stockton M. Mayer, Anwaruddin Syed, Sihai Dave Zhao, Ruoqing Zhu, Rashid Bashir, Nathan I. Shapiro, Bobby Reddy

## Abstract

**BACKGROUND:** Early identification of patients at risk for sepsis, mortality, and clinical deterioration is essential for improving outcomes, but existing diagnostic and predictive tools have limited accuracy. The objective was to evaluate the performance of an FDA-authorized AI tool, the Sepsis ImmunoScore, compared to widely available biomarkers and clinical tools for diagnosis of sepsis and prediction of in-hospital mortality and intensive care unit (ICU) admission.

**METHODS:** This multicenter observational study included 6,027 adult patients suspected of infection across 7 U.S. hospital sites. The Sepsis ImmunoScore’s predictive performance was compared to the sequential organ failure assessment (SOFA) score, procalcitonin (PCT), C-reactive protein (CRP), Systemic Inflammatory Response Syndrome (SIRS) score, National Early Warning Score (NEWS), and quick SOFA (qSOFA). Primary outcomes included sepsis as defined by Sepsis-3 criteria, in-hospital mortality, and ICU admission. Predictive accuracy was assessed using area under the receiver operating characteristic curve (AUC), and 95% confidence intervals were generated and hypothesis testing conducted using the bootstrap method.

**RESULTS:** The Sepsis ImmunoScore demonstrated statistically significant superior performance across all outcomes. For sepsis prediction, the Sepsis ImmunoScore achieved an AUC of 0.82, compared to SOFA (0.72), procalcitonin (PCT) (0.70),C-reactive protein (CRP) (0.61), SIRS (0.59), NEWS (0.69), and qSOFA (0.67). For in-hospital mortality prediction, the Sepsis ImmunoScore achieved an AUC of 0.80, outperforming SOFA (0.72), PCT (0.67), CRP (0.58), SIRS (0.60), NEWS (0.72), and qSOFA (0.69). For ICU admission, the Sepsis ImmunoScore reached an AUC of 0.74, superior to SOFA (0.63), PCT (0.64), CRP (0.54), SIRS (0.60), NEWS (0.70), and qSOFA (0.65). All differences between the Sepsis ImmunoScore and comparators were statistically significant.

**CONCLUSIONS:** The Sepsis ImmunoScore significantly improved predictive accuracy for sepsis, in-hospital mortality, and ICU admission compared to six conventional clinical scores and biomarkers. This AI-based tool may enhance risk stratification and clinical decision-making, potentially leading to more timely sepsis interventions and improved outcomes.

**KEY POINTS:** *Question:* How does the FDA-authorized Sepsis ImmunoScore compare to conventional sepsis tools at diagnosing and predicting sepsis, clinical deterioration, and in-hospital mortality?

*Findings:* In a multicenter observational cohort of 6,027 patients with suspected infection, the Sepsis ImmunoScore demonstrated statistically superior performance compared to PCT, CRP, SOFA, qSOFA, SIRS, and NEWS in predicting all outcomes: sepsis diagnosis, ICU admission, and in-hospital mortality.

*Meaning:* Because the Sepsis ImmunoScore outperforms existing sepsis diagnostics, it could potentially enhance risk stratification and clinical decision-making for patients with suspected infection, enabling more appropriate and timely interventions.

---

**S**epsis is a serious, common, and heterogeneous medical syndrome caused by a dysregulated host response to infection.(1) It is a major cause of morbidity and mortality in the United States and worldwide.(2) Early source control can improve patient outcomes, but prompt recognition of sepsis is challenging.(3) There remains an unmet clinical need for risk assessment tools that enable clinicians to promptly identify patients at risk of sepsis and consequent deleterious outcomes.

Many proposed approaches exist, including laboratory tests, sepsis bio-markers, clinical scores, and artificial intelligence (AI) tools, but none has gained universal acceptance in clinical practice. A recent addition is the Sepsis ImmunoScore™, which became the first AI diagnostic for sepsis authorized by the Food and Drug Administration (FDA) in 2024.(4, 5) It uses machine learning (ML) and up to 22 patient-specific data points, including laboratory measurements, vitals, demographics, and sepsis biomarkers, to predict the risk of sepsis within 24 hours (**Table 1**).

**TABLE 1.** Sepsis ImmunoScore Input Parameters.

| Parameter | Required for Sepsis ImmunoScore result? |
| --- | --- |
| Systolic blood pressure | Yes |
| Diastolic blood pressure | Yes |
| Temperature | Yes |
| Respiratory rate | Yes |
| Heart rate | Yes |
| Blood oxygen saturation | Yes |
| White blood cell count | Yes |
| Platelet count | Yes |
| Blood urea nitrogen | Yes |
| Creatinine | Yes |
| Procalcitonin | Yes |
| C-reactive protein | Yes |
| Age | No |
| Lymphocyte count | No |
| Neutrophil count | No |
| Potassium | No |
| Chloride | No |
| Total carbon dioxide | No |
| Sodium | No |
| Albumin | No |
| Bilirubin | No |
| Lactate | No |

Additional details on data source, time period for valid measures, and selection decisions when multiple measurements are available appear in a prior publication.(4)

The pivotal study also found the Sepsis ImmunoScore to be predictive of other negative outcomes, including in-hospital mortality, length of hospital stay, intensive care unit (ICU) admission, mechanical ventilation, and vasopressor medication use.(4) The approved algorithm is locked to avoid overfitting the model, meaning it is not adjusted for individual institutions, and provides a user-friendly results display explaining a patient’s risk for each outcome and the clinical or laboratory data that weighed most heavily in calculating their score (**Figure 1**).

**Figure 1.**
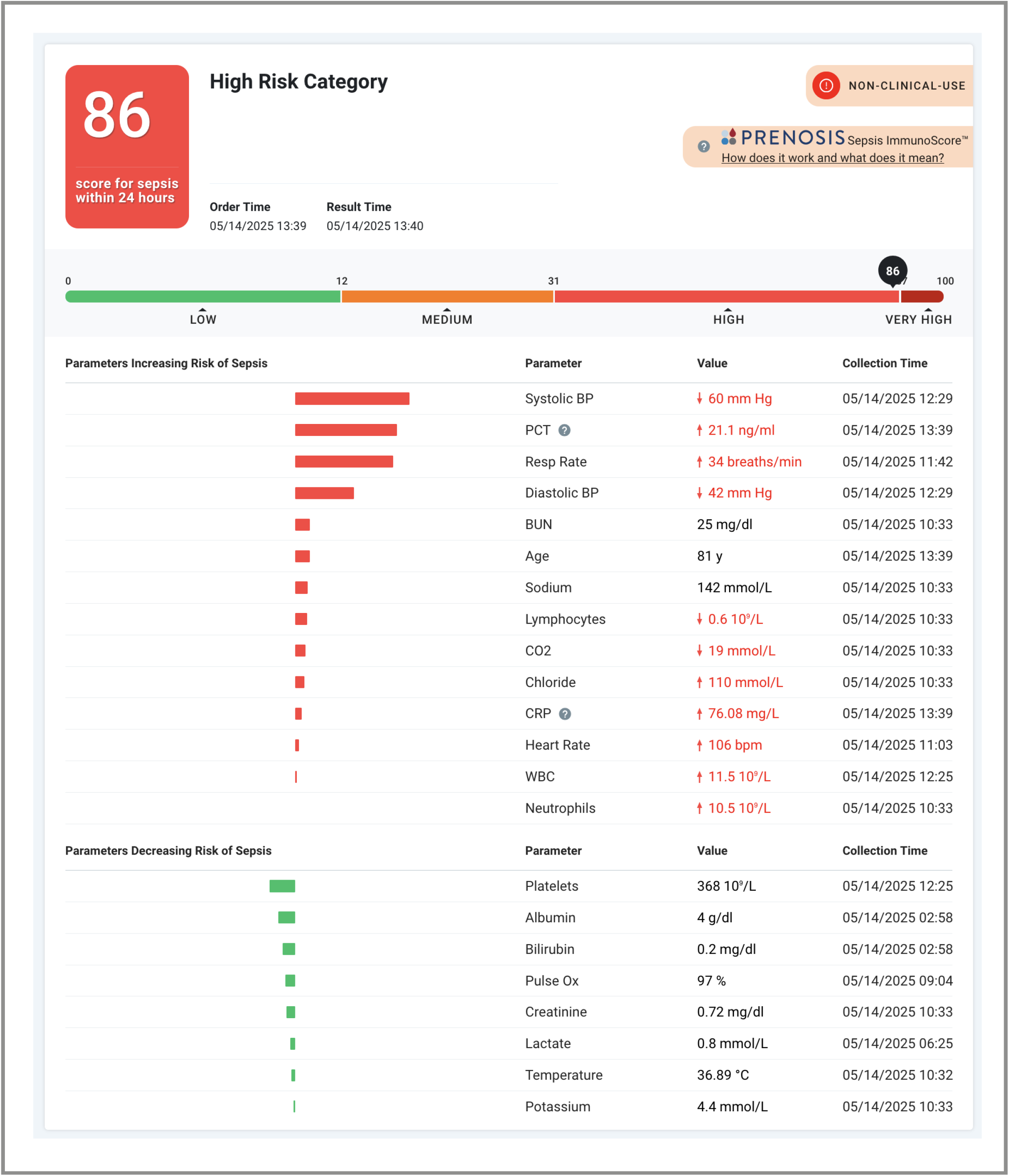
Sample Sepsis ImmunoScore Result Report.

The objective of this study was to compare the predictive performance of the Sepsis ImmunoScore against six salient comparators in a large prospective observational study. We selected the sequential organ failure assessment (SOFA) score, procalcitonin (PCT), C-reactive protein (CRP), the Systemic Inflammatory Response Syndrome (SIRS) score, the National Early Warning Score (NEWS), and the quick SOFA (qSOFA) score for this comparison.

The SOFA score is a clinical score of organ dysfunction that has been used to predict mortality, especially for ICU patients, and is part of the Sepsis-3 definition of sepsis.(1, 6, 7) PCT and CRP are FDA-approved biomarkers of infection that are widely available in clinical laboratories. PCT is a host-response biomarker upregulated by bacterial endotoxins and proinflammatory cytokines, including interleukin-6 and tumor necrosis factor-α. It has been found to be associated with mortality in patients with sepsis and patients with community-acquired pneumonia.(8, 9) CRP is a non-specific marker of inflammation that is associated with mortality in patients with sepsis.(10) SIRS reflects an exaggerated response to infection, surgery, acute inflammation, or another stressor and is evaluated using cutoffs for body temperature, heart rate, respiratory rate, and leukocyte count.(11) A SIRS score of 2 or higher on the first day of hospitalization has been linked to prolonged ICU admission, a greater likelihood of developing multiple organ dysfunction syndrome (MODS), and increased need for vasopressors and mechanical ventilation.(11) NEWS scores include measures such as supplemental oxygen usage, vital signs, and level of consciousness; the measure has demonstrated the ability to predict death, ICU admission, and cardiac arrest within 24 hours and is broadly used for acute care assessments in the United Kingdom and elsewhere.(12) The qSOFA uses any two of its three criteria (Glasgow coma scale, respiratory rate and blood pressure) to identify patients at an increased risk of sepsis, prolonged ICU stay, or in-hospital mortality.(1)

We evaluated the performance of the Sepsis ImmunoScore and these six comparators for predicting sepsis, in-hospital mortality, and admission to the ICU. These tools are often used in triaging patients to determine whether they merit further testing, escalation in therapy, or admission to the ICU. Furthermore, they can be integrated into electronic medical record (EMR) systems to automatically trigger sepsis alerts, which frequently include SIRS and/or SOFA criteria.(13–16) Unlike PCT, CRP, and the Sepsis ImmunoScore, which all must be specifically ordered by a clinician, an EMR-based sepsis alert system can run constantly in the background, generating pop-up notices for all patients meeting the system’s criteria for possible sepsis. While these tools aim to increase early sepsis identification and intervention, they often have very high false-positive rates that result in alert fatigue and even complete dismissal of the warnings by clinicians.(14) Some alert systems show modest success in improving hospital compliance with certain SEP-1 bundle criteria (a set of actions that must be taken within the first 3–6 hours following sepsis identification in order to receive reimbursement from the Centers for Medicare and Medicaid Services), but most automated alerts do not improve mortality outcomes.(14) Other AI-based automated sepsis tools often incorporate criteria from these clinical scoring systems in addition to data such as lab values or even clinical notes. These tools may outperform standard sepsis alerts and even physician diagnosis,(17–20) but as they are proprietary, not FDA-approved, and not widely used (with the exception of the EPIC alert system), they were not included in this analysis.

## MATERIALS AND METHODS

### Study Design

This was a retrospective analysis of prospectively collected blood samples and linked EMR data from a multicenter, observational study of hospitalized patients with suspected infection. Subjects were enrolled at 1 of 7 participating U.S. hospitals between March 2018 and June 2025. The study was approved by the ethics boards of each institution under a waiver of informed consent, except for OSF Saint Francis Medical Center, which required informed consent.

### Study Population

The study enrolled hospitalized adult patients (aged 18 years and older) receiving care in any hospital department with a suspected serious infection, as indicated by a clinician’s decision to order a blood culture. To qualify, participants needed to have a lithium-heparin (Li-Hep) plasma sample drawn and collected and to have a valid Sepsis ImmunoScore result when retrospectively assessed at the time of their first blood culture order. There were no other exclusion criteria. The Sepsis ImmunoScore algorithm was developed using a separate patient cohort;(4) the study population did not include patients on whom the model was trained.

### Data Collection

Remnant blood samples were prospectively collected from all eligible patients as part of an ongoing biobank run by Prenosis, Inc. The biobank performed PCT and CRP testing on the Li-Hep remnants, which were deidentified and assigned unique specimen numbers by a research coordinator prior to transport. Clinical data were extracted from the EMR via direct offline extraction, de-identified, and subsequently linked to biobanked samples. The data transfer did not include protected health information. Extracted data included demographic variables, ICD-10 diagnostic codes, medication records, vital signs, and results from routine laboratory tests such as blood chemistry and lactate levels. Comorbidities were classified according to the Charlson Comorbidity Index, ICD-10-Clinical Modification (CM) codes, and definitions from the National Cancer Institute’s Comorbidity Index and the Surveillance, Epidemiology, and End Results (SEER) program.(21) Immunocompromised status was determined using ICD-10-CM criteria established by the Agency for Healthcare Research and Quality.(22)

Further details on data anonymization, retrieval, classification, and analysis are described in separate publications.(4, 23) A Sepsis ImmunoScore was generated for each patient as an eligibility criterion; SOFA, SIRS, NEWS, and qSOFA scores were generated for each patient retrospectively using EMR data at the time of analysis.

### Study Outcomes

There were three outcomes of interest: sepsis, in-hospital mortality, and transfer to the ICU (in patients not in the ICU when blood cultures were first ordered). Sepsis was assessed using an automated Sepsis-3 label determined retrospectively from each patient’s EMR.(24) In-hospital mortality and ICU admission status were determined from standardized data (Fast Healthcare Interoperability Resources) in the patient EMR.

### Statistical Analysis

Predictive performance was assessed using the area under the receiver operating characteristic curve (AUC). We used 1,000 bootstrapped samples to estimate 95% confidence intervals and conduct hypothesis tests for the difference in AUC between the Sepsis ImmunoScore and each of the comparators.(25) A total of 18 hypothesis tests were conducted. We used the Bonferroni correction to account for multiple testing, rejecting each null hypothesis if the p-value was less than 0.05/18.(26) The analysis was conducted using R statistical software version 4.4.1 (R Foundation for Statistical Computing; Vienna, Austria).

## RESULTS

There were a total of 6,027 patient encounters with valid Sepsis ImmunoScore results during the study period (**Table 2**). Mean age was 64.2 years (SD 17.5 years) with a slight male predominance (52.9%), and two-thirds (67.1%) of the population were White. High-risk comorbidities were common, and more than one-third of the sample (35.2%) met the automated Sepsis-3-based criteria for sepsis within 24 hours of the first ordered blood culture. Mean length of stay was nearly 10 days, and 27.0% of patients were transferred to the ICU, with 6.6% dying during their hospital stay.

**TABLE 2.**
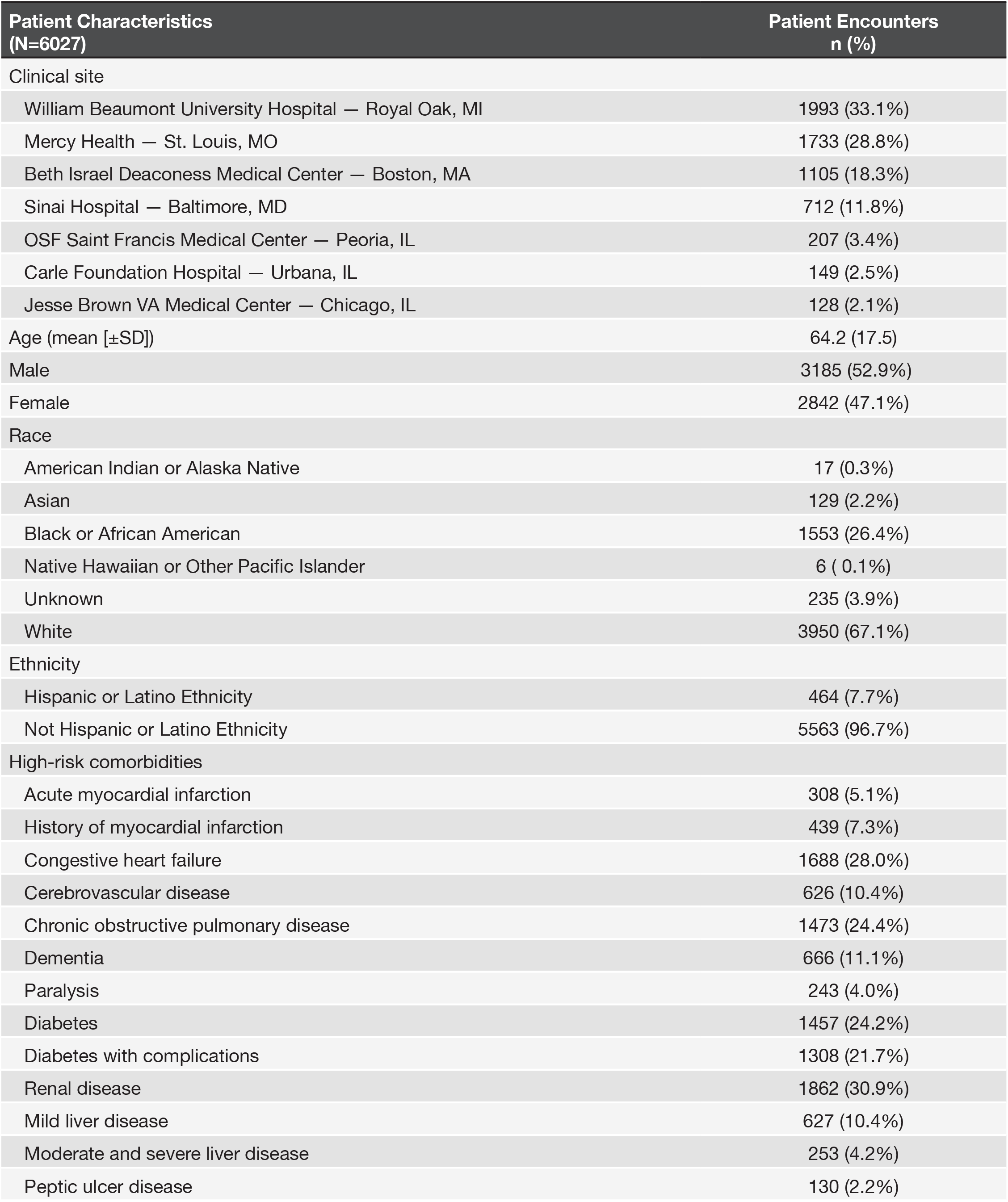
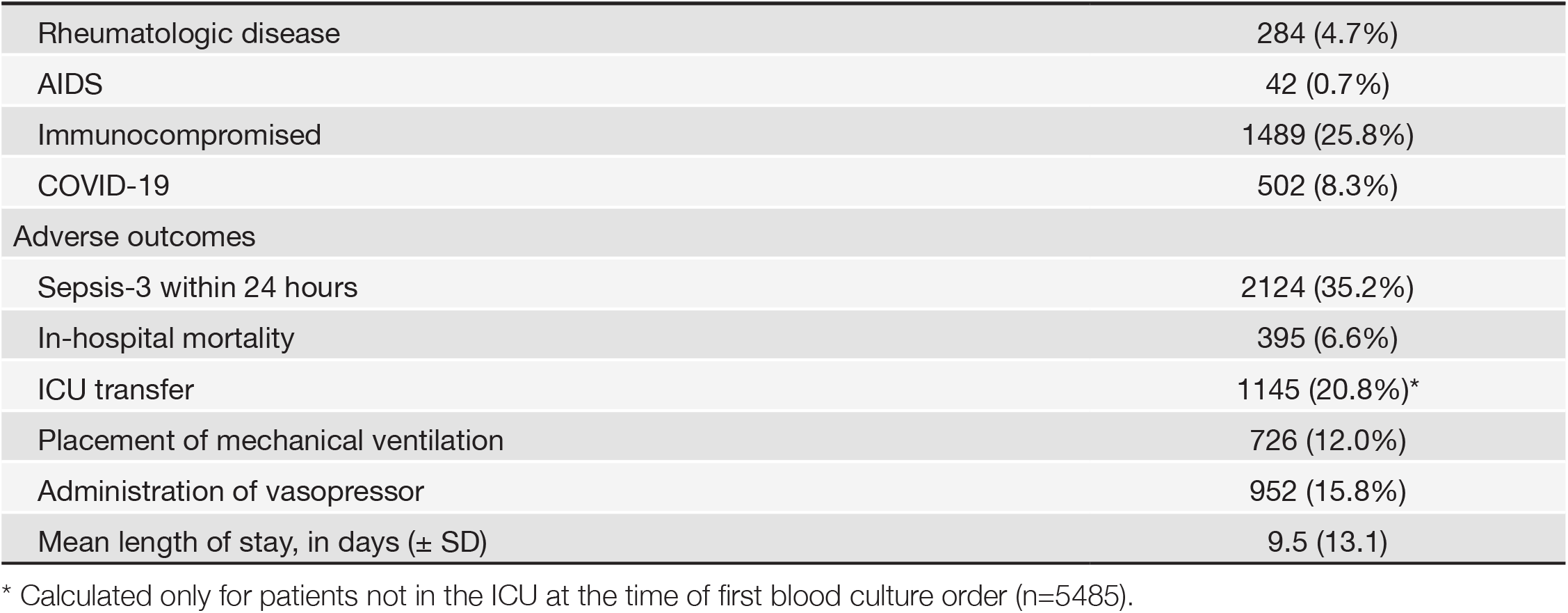
Baseline Data and Adverse Outcomes.

| Patient Characteristics<br>(N=6027) | Patient Encounters<br>n (%) |
| --- | --- |
| Clinical site |  |
| William Beaumont University Hospital — Royal Oak, MI | 1993 (33.1%) |
| Mercy Health — St. Louis, MO | 1733 (28.8%) |
| Beth Israel Deaconess Medical Center — Boston, MA | 1105 (18.3%) |
| Sinai Hospital — Baltimore, MD | 712 (11.8%) |
| OSF Saint Francis Medical Center — Peoria, IL | 207 (3.4%) |
| Carle Foundation Hospital — Urbana, IL | 149 (2.5%) |
| Jesse Brown VA Medical Center — Chicago, IL | 128 (2.1%) |
| Age (mean [±SD]) | 64.2 (17.5) |
| Male | 3185 (52.9%) |
| Female | 2842 (47.1%) |
| Race |  |
| American Indian or Alaska Native | 17 (0.3%) |
| Asian | 129 (2.2%) |
| Black or African American | 1553 (26.4%) |
| Native Hawaiian or Other Pacific Islander | 6 (0.1%) |
| Unknown | 235 (3.9%) |
| White | 3950 (67.1%) |
| Ethnicity |  |
| Hispanic or Latino Ethnicity | 464 (7.7%) |
| Not Hispanic or Latino Ethnicity | 5563 (96.7%) |
| High-risk comorbidities |  |
| Acute myocardial infarction | 308 (5.1%) |
| History of myocardial infarction | 439 (7.3%) |
| Congestive heart failure | 1688 (28.0%) |
| Cerebrovascular disease | 626 (10.4%) |
| Chronic obstructive pulmonary disease | 1473 (24.4%) |
| Dementia | 666 (11.1%) |
| Paralysis | 243 (4.0%) |
| Diabetes | 1457 (24.2%) |
| Diabetes with complications | 1308 (21.7%) |
| Renal disease | 1862 (30.9%) |
| Mild liver disease | 627 (10.4%) |
| Moderate and severe liver disease | 253 (4.2%) |
| Peptic ulcer disease | 130 (2.2%) |
| Rheumatologic disease | 284 (4.7%) |
| AIDS | 42 (0.7%) |
| Immunocompromised | 1489 (25.8%) |
| COVID-19 | 502 (8.3%) |
| Adverse outcomes |  |
| Sepsis-3 within 24 hours | 2124 (35.2%) |
| In-hospital mortality | 395 (6.6%) |
| ICU transfer | 1145 (20.8%)* |
| Placement of mechanical ventilation | 726 (12.0%) |
| Administration of vasopressor | 952 (15.8%) |
| Mean length of stay, in days (± SD) | 9.5 (13.1) |

\* Calculated only for patients not in the ICU at the time of first blood culture order (n=5485).

The AUC of the Sepsis ImmunoScore (0.82; 95% CI: 0.81–0.83) for predicting sepsis within 24 hours of first blood culture order was significantly greater than the AUCs of all six comparators (p < 0.001) (**Figure 2**). Likewise, the AUC of the Sepsis ImmunoScore for in-hospital mortality (0.80; 95% CI: 0.78–0.82) and admission to the ICU (0.74, 95% CI: 0.72–0.75) were both significantly better than those of all comparators (p < 0.001) (**Figure 2**).

**Figure 2.**
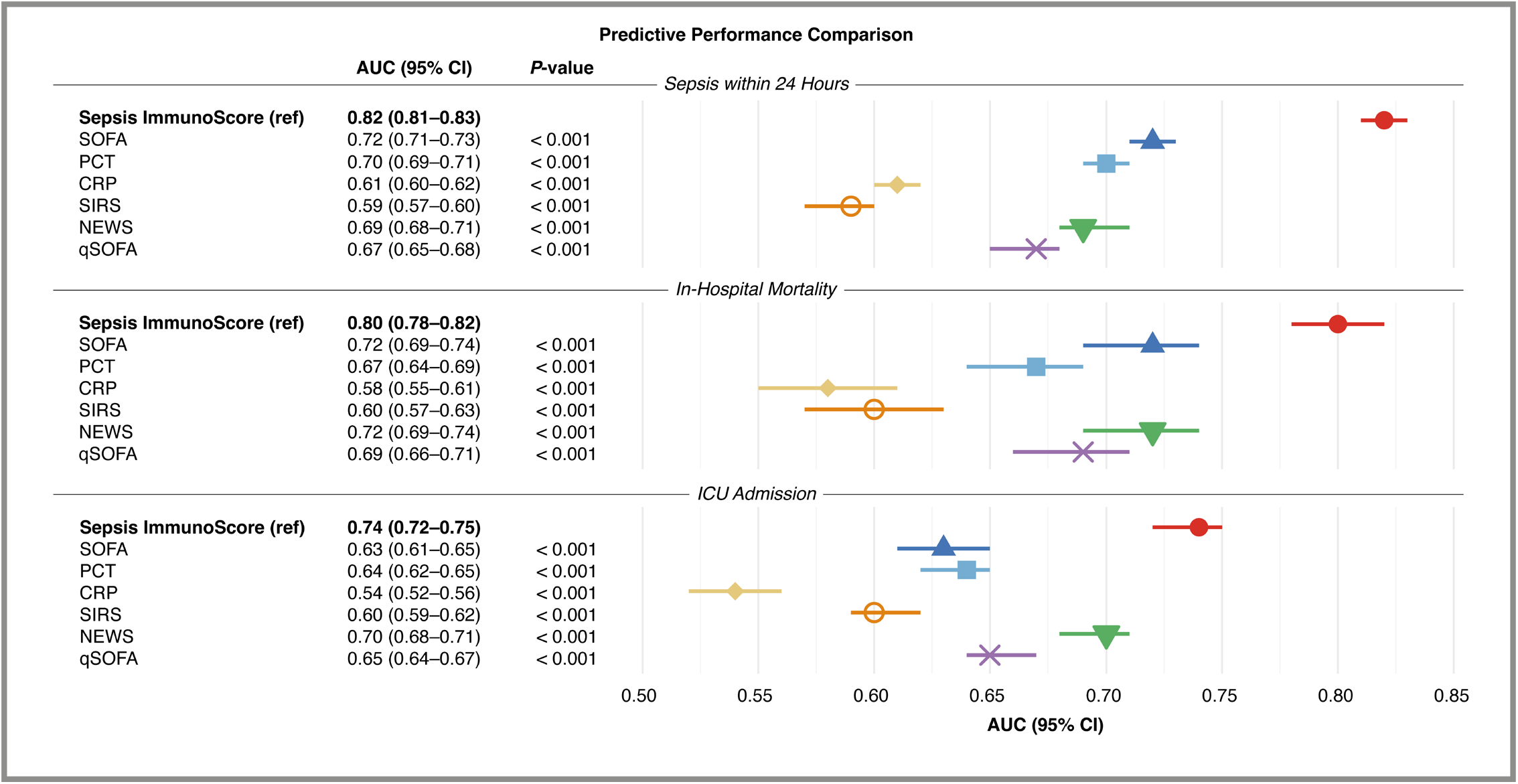
Comparative AUCs of Predictive Tools Across Study Outcomes.

## DISCUSSION

In this large, multicenter study we found that the Sepsis ImmunoScore significantly outperformed six widely available biomarkers and clinical scoring tools for the prediction of sepsis, in-hospital mortality, and clinical deterioration (as indicated by admission to the ICU) for patients suspected of serious infection.

This potentially has several clinically meaningful ramifications.

Delayed recognition of septic shock may delay administration of antimicrobials and interventions that support tissue perfusion (e.g., fluid resuscitation and vasopressors), with a resulting increase in the risk of negative outcomes.(27–30) The connection between rapid administration of broad-spectrum antibiotics and reduced mortality in patients suspected of sepsis has been repeatedly described.(27, 31–33) More accurate predictions may improve risk stratification and prompt more timely interventions, which in turn promise to improve outcomes.

For patients with signs of severe illness, a test result suggesting a low likelihood of sepsis may direct clinician investigation toward alternative explanations for patient symptoms, aiding in differential diagnosis. Improved risk stratification may also make the Sepsis ImmunoScore useful for other clinical decisions, such as patient disposition. Knowing that a patient is unlikely to be septic or to require escalated therapy may, when physician judgment aligns, support the decision to de-escalate therapy, refrain from use of unnecessary antimicrobials, keep a patient on a standard inpatient floor, or even discharge a patient home. This opportunity to prompt further investigations for sick patients, keep higher-level beds open for more at-risk patients, and send healthy patients home sooner could result in more efficient use of healthcare resources, reduced wait times for ICU admission for the sickest patients, better antimicrobial stewardship, and improved patient safety.

Early and more accurate identification of sepsis may have financial impacts as well. Improved diagnosis can aid hospitals in SEP-1 bundle compliance by enabling prompt initiation of the required interventions and assessments. Results from an FDA-approved diagnostic facilitate accurate patient coding and provide documentation for sepsis-related billing and claims. Together, improved SEP-1 compliance and appropriate coding have substantial potential to impact hospital reimbursement for the complex management of sepsis patients.

The Sepsis ImmunoScore differs from other FDA-authorized diagnostic tools for sepsis in its superior performance and holistic assessment of patient risk for sepsis, ICU admission, and in-hospital mortality. Unlike many FDA-approved diagnostic tools for infection that focus on limited markers—such as PCT, CRP, monocyte distribution width,(34) leukocyte biophysical properties,(35) or immune-related gene expression(36)—the Sepsis ImmunoScore integrates multidimensional inputs, including PCT, CRP, vital signs, and clinical data commonly used in scoring systems like SOFA, SIRS, and NEWS. This holistic integration of patient-specific factors enables the Sepsis ImmunoScore to reflect a comprehensive view of patient risk, enhancing the predictive power beyond that of individual markers or scores.

In addition, the Sepsis ImmunoScore differs from other AI algorithms in that it is currently the only one to obtain FDA authorization for sepsis diagnosis and prediction. Unlike passive surveillance tools that continuously monitor EMR data and trigger alerts, many of which employ the clinical scores or biomarkers studied here,(14, 17) the Sepsis ImmunoScore functions as a clinician-ordered diagnostic test; its score places patients into one of four discrete risk categories and is intended for use in conjunction with clinical assessments.(37) Rather than supplant background alerts, we propose that the Sepsis ImmunoScore could complement existing alert systems by reducing false positives and alert fatigue. For example, hospitals could automatically order the Sepsis ImmunoScore for patients flagged by background alerts, using its patient-specific results report to support decision-making regarding disposition and escalation of care.

### Limitations

There are several limitations to this study. First, it is possible that the sample of patients included from the 7 participating hospitals do not reflect patients elsewhere, and we cannot guarantee generalizability of our findings. We attempted to minimize this possibility with bootstrap analysis and the conservative Bonferroni correction to limit the likelihood of spurious findings; the consistent trend of superior performance of the Sepsis ImmunoScore across all outcomes was expected due to the multifaceted, inclusive design of the algorithm. While the automated determination of sepsis assessed in this study has been widely used,(24) it may result in some classification error compared to gold standard chart review. Likewise, missing data from EMRs could have caused misclassification or missed comorbidities.

While we selected widely available, well-known tools and biomarkers for sepsis prediction, we could not compare the Sepsis ImmunoScore to proprietary AI-based algorithms that may have more competitive AUCs for the outcomes of interest. We also did not compare the Sepsis ImmunoScore to passive sepsis alert systems that evaluate every patient regardless of infection-related indicators. By design, the Sepsis ImmunoScore must be specifically ordered by a clinician like any other diagnostic test. This is intended to ensure that sufficient information is obtained to make a reliable assessment of the patient’s risk (via prompts to order any additional tests that the score requires). As such, it cannot identify septic patients for whom the test is not ordered. Instead, it serves two primary purposes: to provide documented support for a practitioner’s judgment about a patient’s status (cases very likely to be or not to be sepsis), or to aid in differential diagnosis and clinical decision-making when the clinical picture is unclear.

## CONCLUSIONS

While sepsis is still a nebulous construct with heterogenous presentations and a variable disease course, the ability of the Sepsis ImmunoScore to predict mortality and objective metrics of clinical deterioration provides evidence that it reflects important underlying biological risks. As ongoing clinical research refines our understanding of sepsis into one of an overarching problem—extreme, dysregulated immune response—with distinct pathophysiological subtypes,(38–40) we believe holistic, biology-based tools will continue to outperform symptom-based tools, isolated biomarkers, and institutional practice-based indicators in sepsis diagnosis. Just as we have seen cancer outcomes improve as the scientific understanding of cancer evolved, refined definitions of sepsis according to subtypes could provide the opportunity to develop and deploy personalized, effective therapies targeting the key biological underpinnings of each patient’s disease.

While we make progress toward that reality, this study provides evidence that the Sepsis ImmunoScore outperforms numerous existing sepsis evaluation tools. It can be easily integrated into the EMR and offers an FDA-authorized diagnostic to determine a patient’s imminent risk of sepsis as well as a prediction of in-hospital mortality, ICU admission, length of stay, and likelihood of requiring mechanical ventilation or vasopressors. Regardless of whether hospitals implement the Sepsis ImmunoScore as a stand-alone diagnostic or paired with background warning systems, our analysis suggests this AI-based algorithm offers excellent diagnostic accuracy for sepsis. By extension, the Sepsis ImmunoScore has the potential to impact clinical practice and hospital resources through more rapid and appropriate triage, informed treatment decisions, opportunities for greater SEP-1 bundle compliance, and improved documentation for coding and claims.

## Data Availability

All data produced in the present study are prioprietary.

## ACKNOWLEDGMENTS

We would like to thank the study coordinators, research staff, and lab technicians for their contributions to this study. Their work was conducted as part of their job duties; they did not receive additional compensation. Writing support was provided by Katherine Brind’Amour of HealthWords, Ltd.

